# Comparison of Tumor Profiles Between Military Personnel and the General Population: Insights from the NHANES Database

**DOI:** 10.1101/2025.04.03.25325223

**Authors:** Cui Pang, Yizhou Luo

## Abstract

**Objective:** This study aims to investigate the impact of military service on tumor inci-dence within the population utilizing data from the NHANES database.

**Methods:** A cross-sectional survey was conducted among individuals aged 18 years and older, as recorded in the NHANES database from 1999 to 2018. The primary outcome measure was the prevalence of self-reported cancer. Multivariate logistic regression analyses were employed to evaluate the association between military service, various covariates, and cancer prevalence.

**Results:** The study included a total of 12,174 participants, of whom 1,904 (15.64%) reported a history of military service. Participants with a history of military service were, on average, older than those without such a history (66.17 ± 0.43 VS 52.79 ± 0.31). Additionally, individuals with military service were more frequently identified as white (85.45% vs. 69.74%, p < 0.001), exhibited higher rates of marriage and annual household income (p < 0.001), and were more likely to be smokers (66.01% vs. 43.32%, p < 0.001). In this study, the observed prevalence of cancer within the population was notably higher than that typically reported in the general population. This dis-crepancy is attributed to the exclusion of individuals who were unaware of their cancer status. The prevalence of cancer was particularly elevated among individuals with a history of military service, with rates of 76.95% compared to 46.87% (p < 0.001). Notably, cancers affecting the reproductive endocrine system, dermatological, skeletal, and other bodily systems were more frequently observed. After adjusting for all available con-founding variables, multivariate analyses revealed that the odds ratio (OR) for having a tumor in individuals without military service, compared to those with military service, was 0.26 (95% CI 0.23-0.30) and 0.40 (95% CI 0.31-0.53), respectively, indicating a signif-icantly reduced likelihood of tumor occurrence.

**Conclusion:** Military service is associ-ated with an increased prevalence of tumors, particularly thyroid cancer. Further re-search is warranted to elucidate the relationship between military service and cancer diagnoses, as well as to identify the underlying causes of this association. Such efforts are essential for enhancing health protection measures for military personnel and re-ducing the incidence of cancer within this population.

## Introduction

Military service is a mandatory duty for citizens in many countries. During their service, military personnel may encounter various carcinogenic risk factors, including exposure to radioactivity [1], ultraviolet radiation [2], chemicals [3], neurotoxicants [4], and radioactive substances [5], at levels higher than those experienced by the general population. Cohort studies involving male Vietnam veterans have demonstrated that exposure to Agent Orange is associated with an increased risk of bladder cancer [6]. Additionally, research indicates that military service members may have a higher propensity for smoking and alcohol abuse, further elevating their cancer risk [7,8]. Despite the availability of a specialized, free healthcare system for military personnel, existing literature suggests that U.S. military service is linked to a higher incidence of cancer, including thyroid [9], breast, and prostate cancer [10], melanoma [11], colorectal cancer [12], and mesothelioma [13].

However, a retrospective study with a 48-year follow-up period found that veterans involved in chemical agent research at Porton Down exhibited a slightly higher mortality rate, but no difference in cancer incidence [14]. In a cohort study with an 11-year follow-up period, British Gulf War veterans exhibited no increased overall cancer risk, nor any site-specific cancer risk [15]. Additionally, research has indicated that the incidence of neuroepithelial brain cancer [16], digestive system cancers[17], bladder and kidney cancers [18], soft-tissue sarcomas [19], and oral cavity and oropharyngeal cancers [20] is lower among active-duty military personnel compared to the general U.S. population.

To further explore the disparities in cancer incidence and the spectrum of cancer types between military service members and the general population, we utilized the NHANES database. This analysis adjusted for various demographic and socioeconomic factors to examine cancer prevalence among individuals with and without a history of U.S. military service.

## Methods

### 2.1 Study design and subjects

Following the STROBE Statement [21], the cross-sectional study utilized NHANES data covering the period from 1999 to 2018. The National Health and Nutrition Examination Survey (NHANES), conducted by the Centers for Disease Control and Prevention (CDC), is designed to evaluate the health and nutritional status of American participants through a combination of interviews, physical examinations, and laboratory analyses. Comprehensive details regarding the project’s design, data collection methodologies, sample weighting, and informed consent procedures are available through the National Center for Health Statistics (NCHS), with pertinent data being publicly accessible [22]. The data, having been previously anonymized and released by NCHS, do not require Institutional Review Board approval.

### 2.2 Criteria for inclusion and exclusion

Given that military service typically commences at the age of 18, the study’s inclusion criteria were restricted to individuals aged 18 and older. The initial cohort included 119,555 participants, excluding those who either refused to disclose their military service status (N=19), were uncertain about their service status (N=7), had missing data regarding service status (N=55,923), were under 18 years of age (N=2,153), refused to disclose their cancer history (N=3), were uncertain about their cancer history (N=57), had missing cancer history data (N=4,180), with 3 or more tumors (N=14), refused to answer tumor type (N=42), did not know tumor type (N=92), had missing tumor type data (N=44890), or had incorrect data (N=1). In total, 12,174 participants were included in this cross-sectional analysis. The process for selecting participants is detailed in Figure 1.

**Fig 1.**
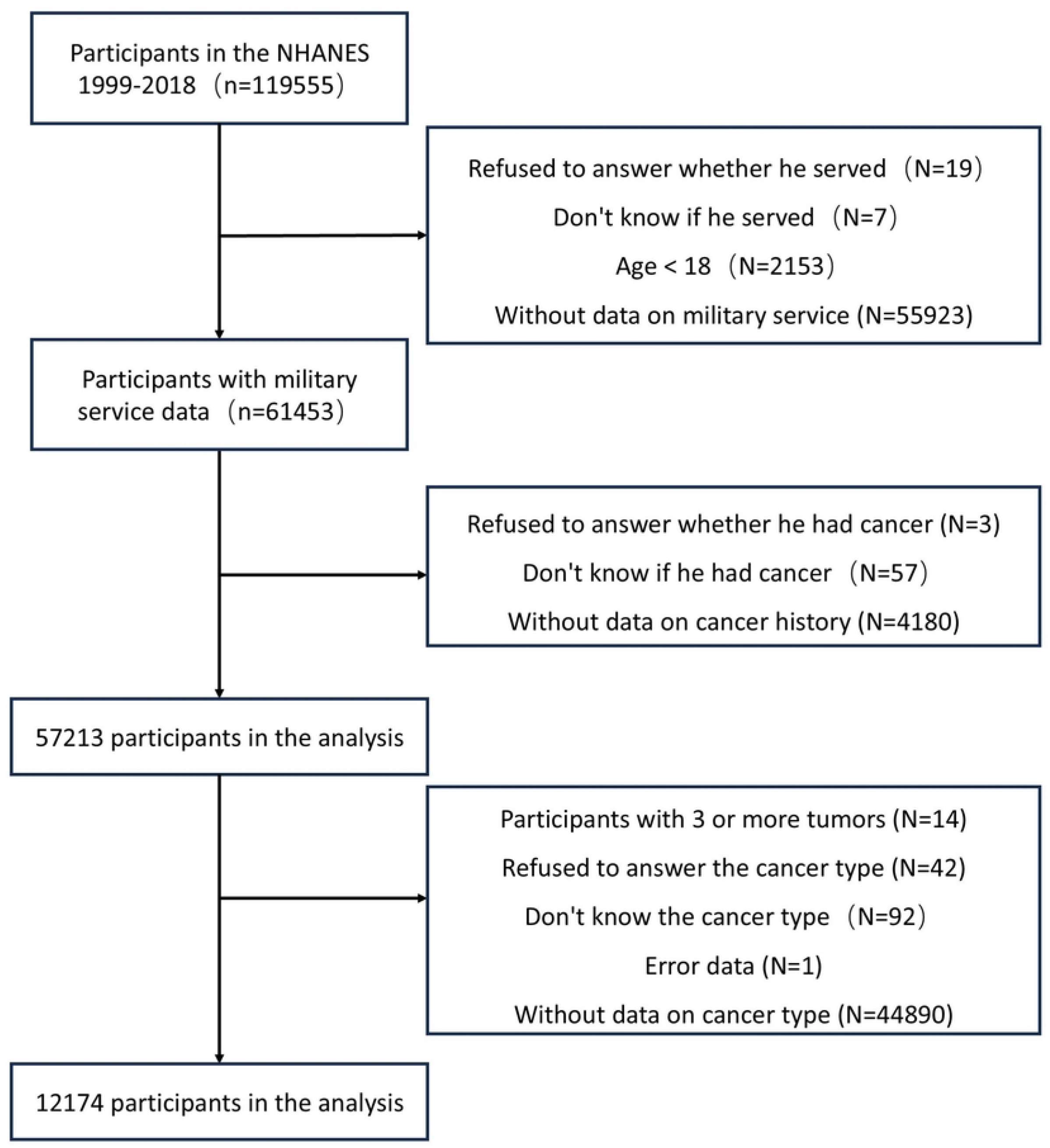
Participant Censoring Flowchart.

### 2.3 Endpoints and exposure factors

The primary endpoint of this study was the self-reported incidence of any cancer diagnosis, assessed using the survey question associated with the variable DMQMILIAT, which inquired whether the respondent had served in the U.S. Armed Forces. The survey question linked to the variable MCQ220 was employed to ascertain whether the respondent had ever been informed by a doctor or other medical professional of having any form of cancer or malignancy. Furthermore, the survey question with the variable name MCQ230A was utilized to identify the specific type of cancer diagnosed. The cancer types were subsequently categorized according to anatomical systems. Tumors of the urinary system included bladder and kidney cancers; tumors of the hematological system encompassed hematological tumors, leukemia, and Hodgkin’s lymphoma; tumors of the digestive system comprised colon, esophageal, gallbladder, liver, pancreatic, rectal, and gastric cancers; and tumors of the reproductive and endocrine systems included breast, cervical, testicular, uterine, prostate, thyroid and ovarian cancers; tumors of the nervous system included brain tumors and other nervous system tumors; tumors of the skin and soft tissue encompassed melanoma, skin cancer, soft tissue tumors, as well as cancers of the mouth, tongue, and lip; tumors of respiratory system comprised laryngeal, tracheal, and lung cancers; tumors of bone and other types were categorized as bone tumors and miscellaneous tumors.

### 2.4 Covariate

To strengthen the analysis of the relationship between tumors and military service, we incorporated the following variables as covariates: age, gender, race, marital status, education level [23], poverty-to-income ratio (PIR), smoking status [24], alcohol consumption patterns [25, 26], exercise equivalents [27], diabetes, hypertension, and depression scores [28, 29]. Diabetes mellitus was identified based on one or more of the following criteria: a history of diabetes mellitus, use of insulin, use of medication to lower blood glucose, hemoglobin A1c level of 6.5% or higher, fasting blood glucose level of 126 mg/dL or higher, or a 2-hour postprandial blood glucose level of 200 mg/dL or higher. Hypertension was defined as a history of hypertension or having a systolic blood pressure (SBP) greater than 140 mmHg or a diastolic blood pressure (DBP) greater than 90 mmHg.

### 2.5 Statistical analysis

In this study, the test weights recommended by the CDC guidelines were utilized [30-33]. Statistical analyses were conducted using R version 4.3.0. Normally distributed continuous data were presented as mean (standard error), and comparisons between two independent groups were performed using the t-test. Categorical data were expressed as frequencies and percentages (n [%]) and were compared using the chi-square test or Fisher’s exact test. The relationship between military service and cancer incidence was examined using logistic regression models. All analyses were conducted using the R^2^ statistical package (R Foundation). A two-tailed p-value of less than 0.05 was considered indicative of statistical significance.

## Results

The study comprised a total of 12,174 participants, of whom 1,904 (15.64%) were engaged in military service, while 10,270 (84.36%) were not. The mean age for participants in military service was 66.17 ± 0.43 years, compared to 52.79 ± 0.31 years for those not in military service, indicating a statistically significant difference in mean age (t = −27.52, P < .001).

Statistically significant differences were also observed between the two groups in terms of gender, race, and culture, as detailed in Table 1. Furthermore, significant disparities were identified in age, gender, race, culture, marital status, glance, alcohol consumption, hypertension, diabetes, and depression (P < 0.05), whereas no significant differences were found in education and race (P > 0.05).

**Table 1.**
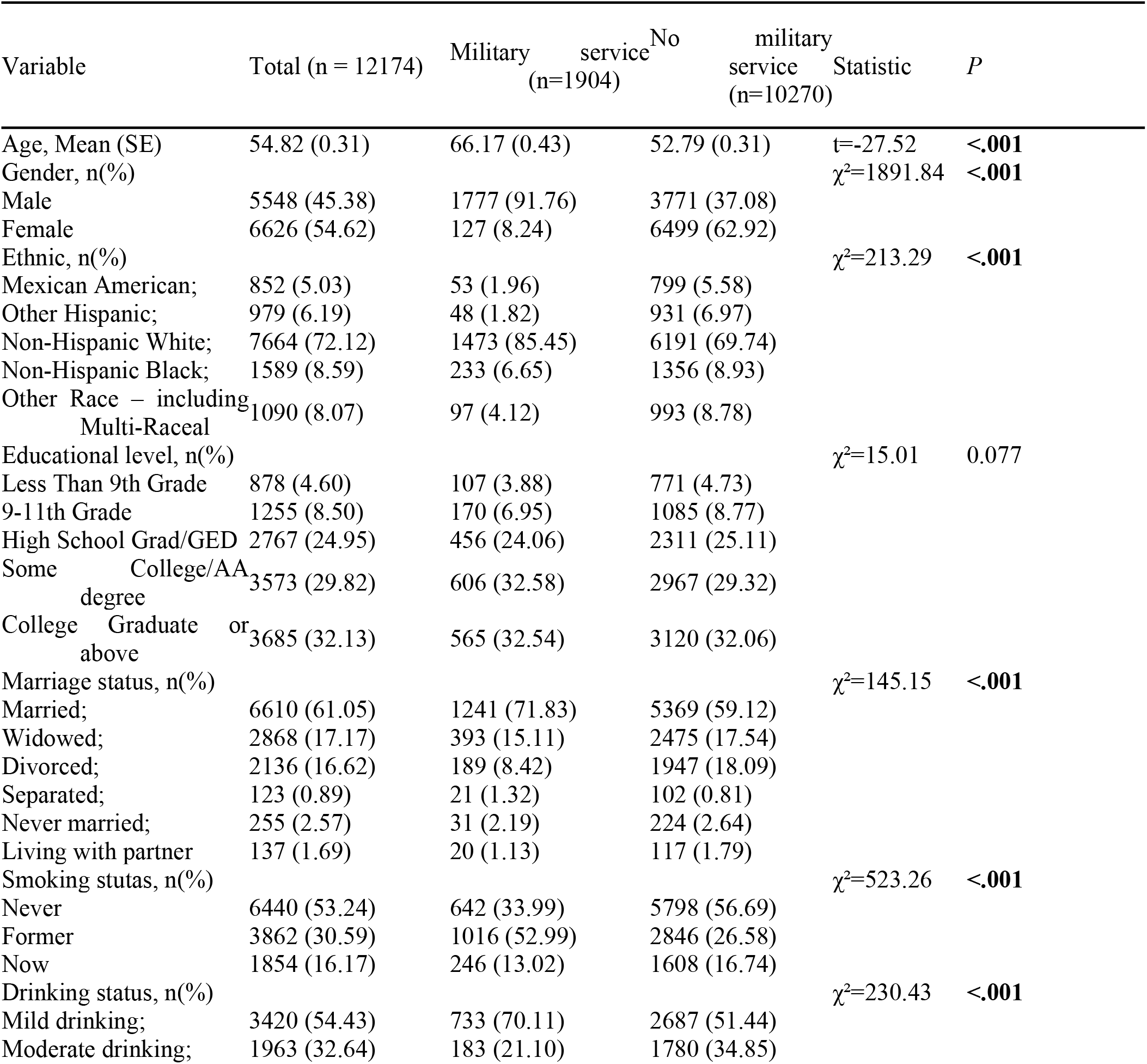
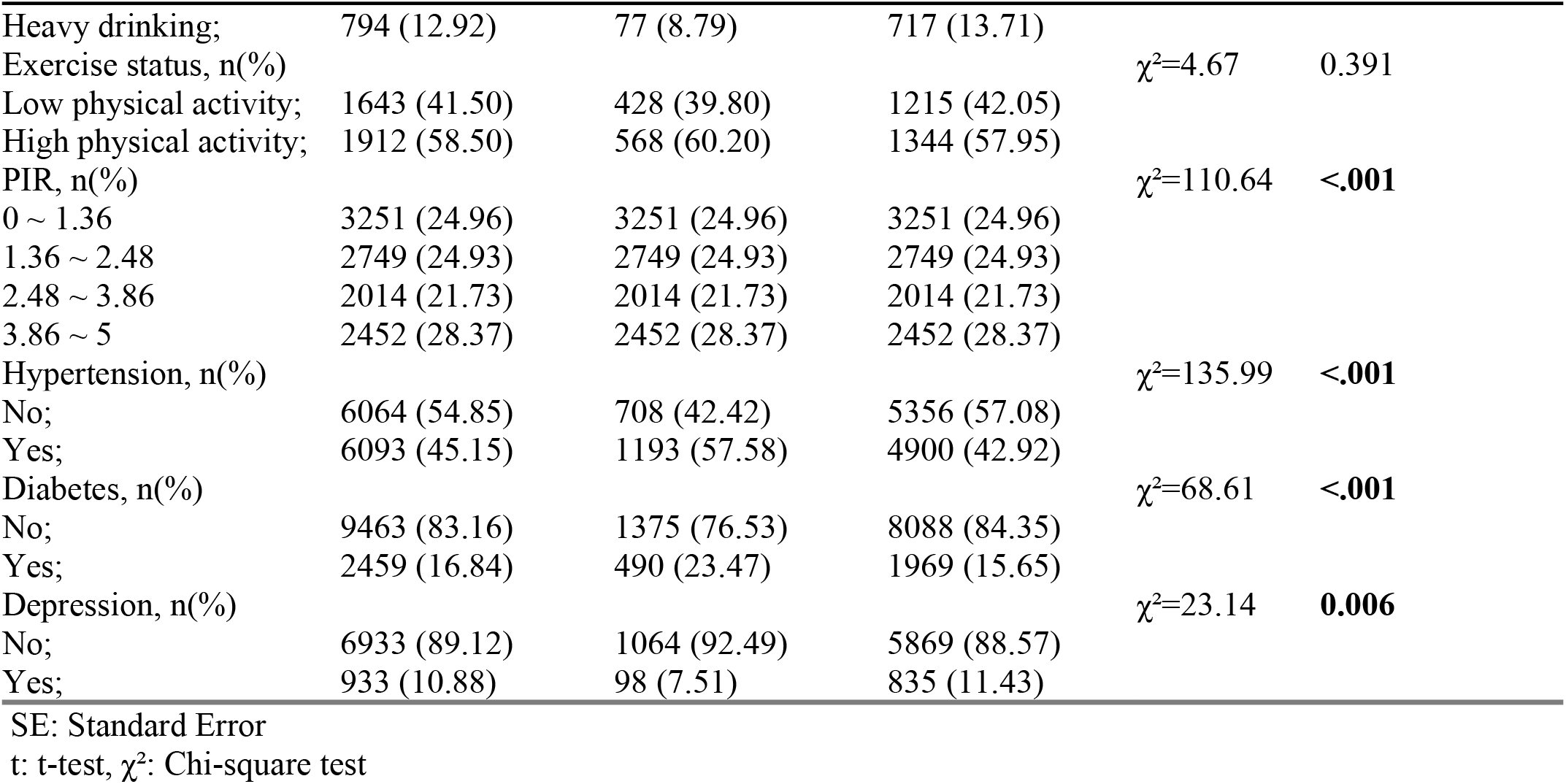
A comprehensive overview of the demographic, socio-economic, and health-related characteristics of the study population.

Figure 2 illustrates the age-stratified proportions of cancer incidence in both military and non-military groups. As depicted in Figure 2, the prevalence of cancer was notably higher in the military group compared to the non-military group, with all cancer types accounting for 1,394 out of 1,904 cases (76.95%) in the military group and 4,162 out of 10,270 cases (46.87%) in the non-military group. In the military cohort, reproductive endocrine tumors were the most prevalent malignant neoplasms, constituting approximately 41.9% of cases, with thyroid cancer being the most frequent at 436 out of 1,394 cases (19.43%). This was followed by bone and other tumors (25.37%), and skin and soft tissue tumors (8.65%). Conversely, in the non-military cohort, digestive system tumors were the most prevalent, accounting for approximately 12.32% of cases, with liver cancer being the most common within this category at 895 out of 10,270 cases (9.10%). This was followed by bone and other tumors (12.15%), and reproductive and endocrine tumors (7.94%). Detailed cancer incidence rates for each type of cancer in both cohorts are provided in Supplementary Table 1, while Supplementary Table 2 presents the incidence rates by anatomical system.

**Table 2.**
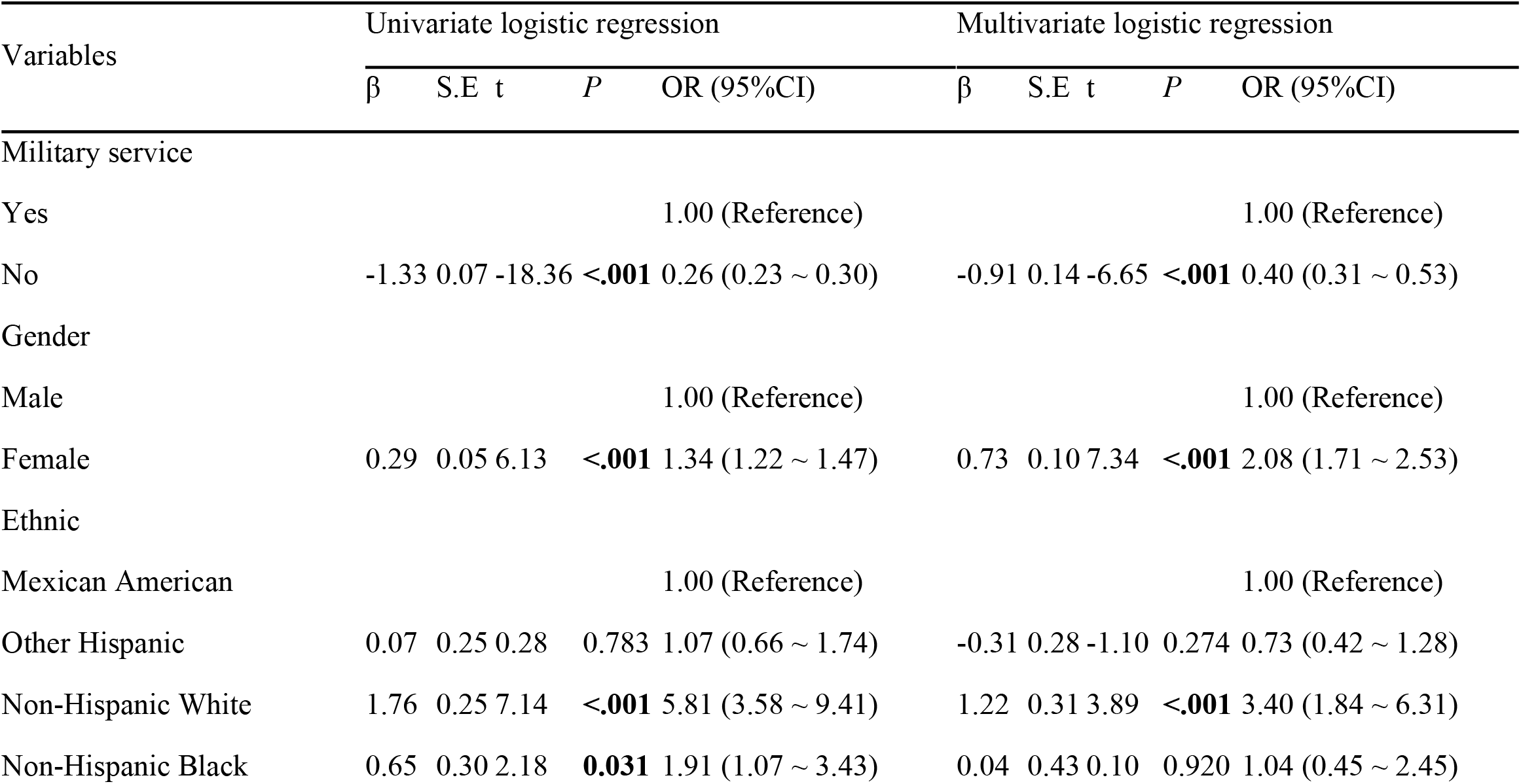
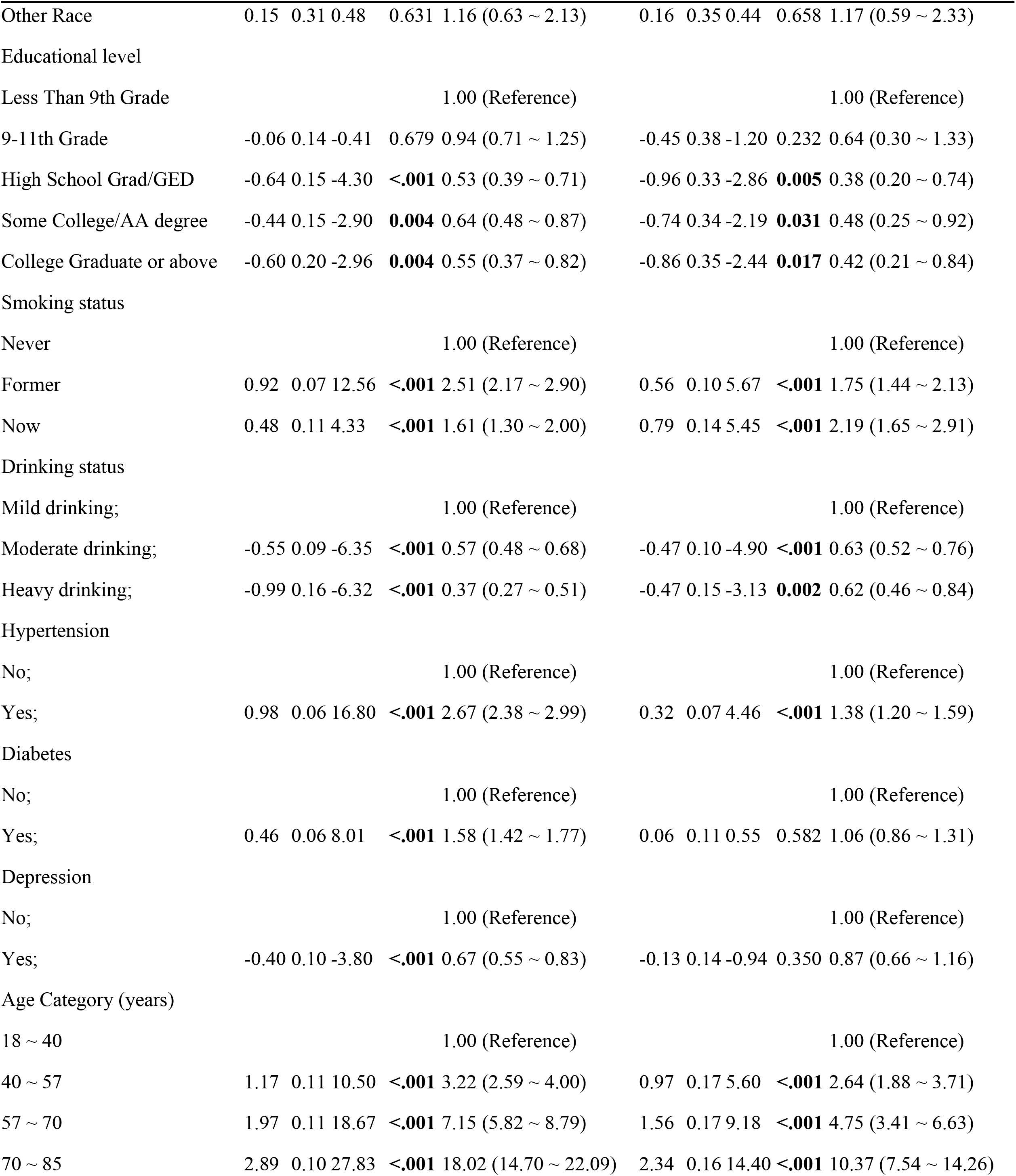
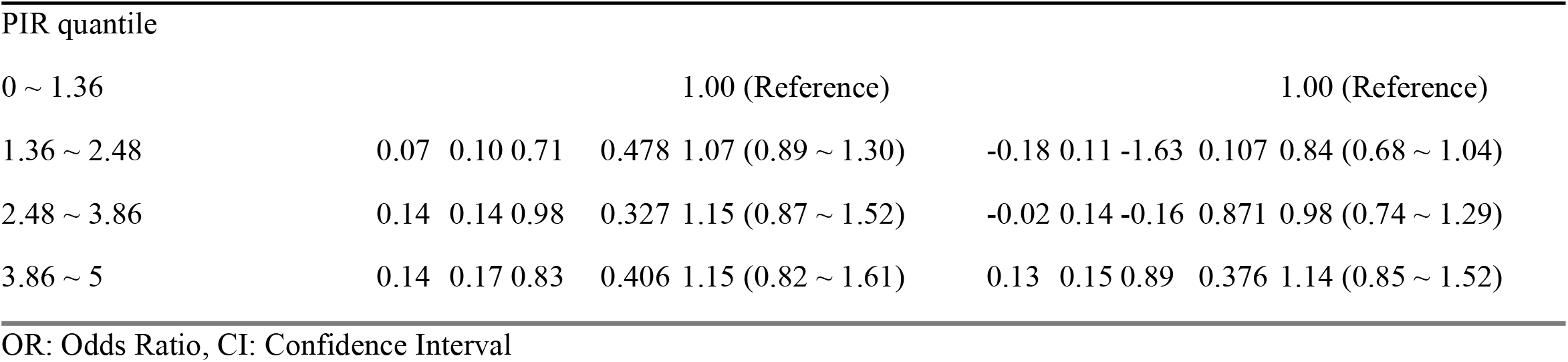
Univariate logistic regression and multivariate logistic regression models showing the association of various covariates with cancer occurrence.

**Fig 2.**
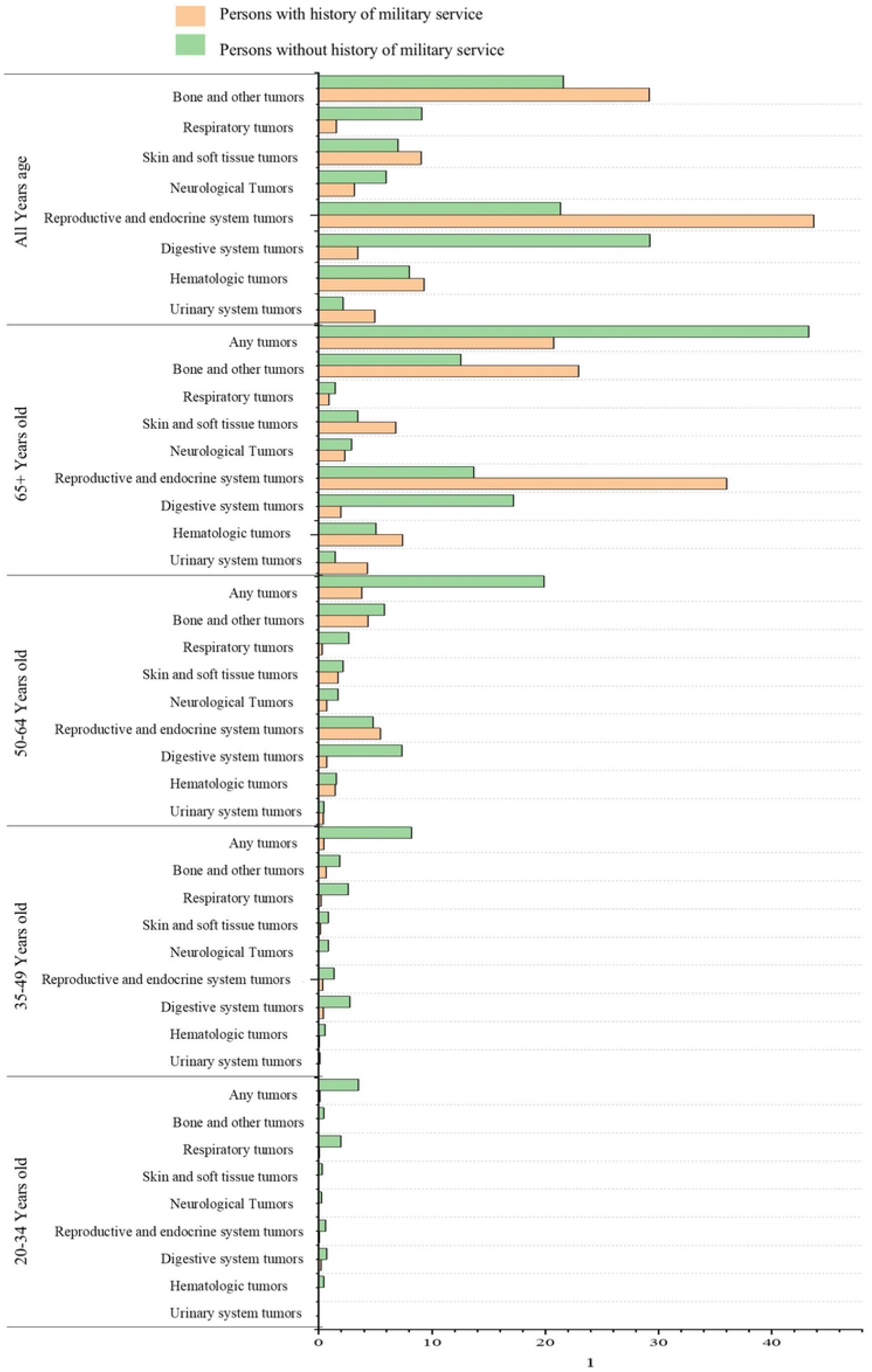
The age-stratified comparison of cancer prevalence between individuals with and without a history of U.S. military service.

Univariate logistic regression and multivariate logistic regression models were employed to evaluate the association between various covariates and cancer occurrence, revealing that the odds ratio (OR) for cancer in the non-military population relative to the military population was 0.4 (95%CI 0.31 ~ 0.53), (p < 0.0001). The likelihood of developing cancer within the population exhibits a significant increase with advancing age. Furthermore, the probability of receiving a cancer diagnosis is notably higher among females compared to males, smokers compared to non-smokers, and Non-Hispanic Whites compared to Mexican Americans (Specific results are detailed in Table 2).

## Discussion

In this study utilizing data from the NHANES database, approximately 15.64% of participants reported a history of military service. Individuals with military backgrounds were predominantly Non-Hispanic White (85.45%), older, more educated, had higher annual household incomes, and exhibited a higher likelihood of smoking (66.01%). The military cohort demonstrated an increased propensity for cancer development within the overall population. Multivariate analysis revealed that a history of military service was significantly associated with a higher prevalence of cancers, excluding those of the digestive and respiratory systems. Furthermore, factors such as age, female gender, and smoking were found to increase the likelihood of cancer, whereas annual household income, obesity, and depression were not significantly correlated with cancer incidence across different systems.

Several plausible explanations exist for the strong association between military service history and cancer diagnosis.

Firstly, individuals with military service tend to be older, which correlates with a higher cancer risk. Secondly, a higher proportion of military personnel have a history of smoking, a well-established risk factor for various cancers, including those of the lung, liver, bladder, kidney. Third, military service exposes personnel to potential carcinogens, including ultraviolet light and radiation. Additionally, the military health system, as the fourth largest in the United States, provides military personnel with organized, accessible, and cost-free healthcare. This may result in more effective screening programs and higher adherence rates, potentially leading to increased cancer diagnoses.

Our study also indicates that individuals with a military background are significantly more likely to develop tumors in the reproductive and endocrine systems, primarily thyroid cancer. This finding aligns with an Italian study, which reported that the standardized incidence of thyroid cancer among Army servicemen is one to two times higher than expected [34]. The etiology of thyroid cancer remains poorly understood, with radiation exposure being the only major recognized risk factor.

Military personnel may experience increased radiation exposure, particularly from ammunition and depleted uranium used in tanks. Furthermore, military personnel are significantly more likely to be exposed to environmental chemicals such as polychlorinated biphenyls (PCBs), polybrominated diphenyl ethers (PBDEs). Extensive nested case-control studies conducted among U.S. military personnel have demonstrated significant associations and dose-response relationships between exposure to specific PCBs and their congeners and the risk of thyroid cancer [35]. Additionally, various endocrine-disrupting chemicals (EDCs), including flame retardants, PCBs, phthalates, and certain pesticides, have been identified as being associated with an elevated risk of thyroid cancer [36].

Our multivariate analysis revealed that factors not related to military service history, such as age, were associated with increased cancer prevalence [37]. Aging has been identified as the most significant risk factor for cancer, as it is associated with the accumulation of cellular damage [38], a decline in immune system function [39], and an increase in chronic inflammation [40]. Consequently, the incidence of most cancers rises substantially with age. The study revealed that 67.9% of veterans exhibited greater longevity compared to the average lifespan of their sex-specific birth cohort. A trend toward a significant increase in longevity over time was observed among veteran males compared to reference males (P < 0.002), whereas no significant trend was detected among females [41]. The average age of military service members was notably higher than that of non-service members, potentially introducing bias into the study.

Several important limitations should be acknowledged. Firstly, the retrospective nature of the data collection may have led to inaccuracies or omissions in data entry. Additionally, the cross-sectional design of the study and the absence of data on critical potential confounders precluded the attribution of causality between military service and cancer occurrence, allowing only for the examination of correlations. Furthermore, although the study accounted for numerous socioeconomic and clinical factors, it did not include established risk factors for malignancy, such as family history, personal genetic predispositions, detailed comorbidity histories, and dietary habits. Despite these limitations, our study constitutes a substantial and distinctive real-world cohort of the American population. Utilizing self-reported data, we identified a history of military service as a potential risk factor for malignancy.

## Conclusion

Individuals with a history of military service may exhibit an elevated risk of cancer, with a higher incidence of thyroid cancer, bone tumors, and skin cancer. This increased risk is likely attributable to improved access to screening and healthcare programs, although it may also be influenced by exposures unique to military environments. The findings of this study underscore the need for prospective research to more accurately assess cancer incidence rates, investigate risk factors for cancer development, and enhance health protection measures for military populations.

## Data Availability

All relevant data are within the manuscript and its Supporting Information files.

## Acknowledgments

Not applicable.

## Data availability statement

All raw data and code are available upon request.

## Ethics statement

Not applicable.

